# Direct Retinal Imaging for Shock Resuscitation in Critical Ill Adults II (D-RISC II)

**DOI:** 10.1101/2025.01.29.25321350

**Authors:** George Cooper, Jamie Burke, Charlene Hamid, Emily Godden, Neeraj Dhaun, Stuart King, Thomas J. MacGillivray, J. Kenneth Baillie, David M. Griffith, Ian J.C. MacCormick

**Author notes:** Corresponding author Corresponding author email address, Corresponding author postal address: *Robert O Curle Ophthalmology Suite, Institute for Regeneration and Repair, University of Edinburgh, UK, EH16 4UU.

## Abstract

**Background:** Shock involves microcirculatory dysfunction that is not suitably captured well by measurements of large vessels, such as systemic blood pressure. The outer retinal microcirculation (the choroid) can be measured non-invasively and may reflect dysfunction in other organs. We tested the feasibility of measuring the retinal choroid in an intensive care setting and explored associations between choroidal measurements and severity of disease.

**Methods:** We performed optical coherence tomography on patients admitted to the intensive treatment unit, and repeated imaging once 12-72 hours later. We measured choroidal anatomy using automated image segmentation, compared this to routine clinical data, and described change over time.

**Results:** Of fifteen patients recruited, 80% (12) had successful baseline imaging and 40% (6) of these had follow-up imaging within intensive care. At baseline, patients with thicker choroids and larger vascularity had larger cumulative fluid balance, and lower disease severity (Acute Physiology and Chronic Health Evaluation II) score, haematocrit, and albumin. A measurable suprachoroidal space was seen in 75% (9) patients and the size of this space tended to be larger in patients with lower heart rates. There was substantial intraindividual variation in choroidal measurements over time.

**Comment:** Measuring the retinal choroid is feasible in patients with critical illness. Exploratory associations with systemic variables suggest that the choroid may provide information about the microvascular function of other major organs. Size and change of choroidal measurements may reflect perfusion pressure or vascular leak in response to inflammation.

## Introduction

Circulatory shock is a life-threatening failure of central organ perfusion, and affects an estimated 30% of patients in intensive care.^1^ Current management aims to improve perfusion by optimizing cardiac output, peripheral vascular resistance, and circulating volume. These are effective at restoring macro-scale physiological measurements, such as blood pressure. However, microvascular function is not routinely monitored because of a lack of reliable, reproducible and practical tools to measure it. In animal models, microvascular function within central organs is directly relevant to perfusion.^2^ The failure of large vessel metrics to reflect microcirculatory perfusion may explain why early goal-directed therapies to restore large vessel parameters appear to be ineffective,^3^ or harmful.^4^ Consistent with this, microvascular perfusion of sublingual capillaries appears to be a better predictor of multiorgan failure in haemorrhagic shock than serum lactate or systolic blood pressure.^5^ However, it is difficult to reproducibly measure the microvascular perfusion of central organs consistently in humans. The retina is an exception, since retinal anatomy provides consistent landmarks, and exposes the central nervous system to direct, non-invasive optical imaging.^6^

The retina is supplied by two vascular networks. The inner retinal circulation perfuses the superficial two-thirds of the retina and is directly visible on ophthalmoscopy. It has a similar blood-tissue barrier, blood flow, and oxygen extraction to the brain.^7^ The outer retina is perfused indirectly by the choroid and choriocapillaris across the outer blood-retina barrier (the retinal pigment epithelium). In contrast to the inner retinal and cerebral vasculature, endothelial cells of the choriocapillaris are fenestrated and choroidal blood flow is ten times that of the brain. The choroidal circulation has parallels with that of the renal cortex.^8^ The choroid can be imaged using commercially available optical coherence tomography (OCT) by using enhanced-depth imaging

(EDI) to penetrate the retinal pigment epithelium. OCT has been used to image the retina in patients in an intensive care setting,^6,9^ but to our knowledge there are no published data describing the choroid in patients being treated for shock. Since choroidal anatomy adapts dynamically to systemic physiology,^10^ and may reflect the perfusion of other central organs,^8^ we aimed to test the feasibility of measuring the choroid with EDI-OCT in patients with shock and explore associations between the choroid and relevant clinical markers.

## Methods

### Participants

We recruited to the Direct Retinal Imaging for Shock Resuscitation in Critical Ill Adults II (D-RISC II) study between March 2023 and July 2023, from the intensive treatment unit (ITU) and high-dependency units (HDU), Royal Infirmary of Edinburgh. All patients admitted during the recruitment period were screened for eligibility on admission by departmental research nurses. Patients were eligible if they were age 16 or older, and receiving Intensive Care Society Level 2 or 3 care. Patients were excluded if they had anticipated survival <24 hours, were pregnant, had obvious bilateral ocular pathology or facial trauma precluding imaging, clinical contraindications to pupil dilation, if the study was likely to disrupt their care, if OCT equipment was not available, or if consent was not given.

### Image capture

We performed baseline EDI-OCT as soon as possible once recruited, and follow-up imaging 12-72 hours later, using the Heidelberg Spectralis Flex (Heidelberg Engineering, Germany) (protocols in Supplement 1), which is mounted on a boom to facilitate retinal imaging of supine patients. This was operated by two people: one to position the camera, and one to operate the computer. In some cases, a third person helped to stabilize the head and eyelids. We recorded systemic data prospectively from clinical records (full variable list in Supplement 2). Imaging was acquired done after pharmacological pupil dilation (Tropicamide 1%). Pupil dilation within a shallow ocular anterior chamber can cause acute angle closure, so anterior chamber depth was tested before dilation.^11^ Follow-up imaging was only performed within ITU/HDU. We aimed to recruit 15 participants to assess feasibility of choroidal imaging.

### Image analysis

The choroid was measured within a 4 mm, horizontal line region of interest centered on the fovea (Figure 1A, top green line). The choroid is defined as the space between the hyperreflective Bruch’s membrane (anterior) and sclera (posterior) (Figure 1A, bottom). The suprachoroid is a potential space between the choroid and the sclera (Figure 1B, top). The choroid is a complex vascular mesh, and the ratio of vascular spaces to whole choroidal space is defined by the choroidal vascular index (CVI) (Figure 1B, bottom red : blue). We measured these features using published open-source automated or semi-automated tools: DeepGPET (choroid),^12^ GPET (suprachoroid),^13^ and MMCQ (choroidal vessel spaces).^14^ Details are in Supplement 3.

**Figure 1:**
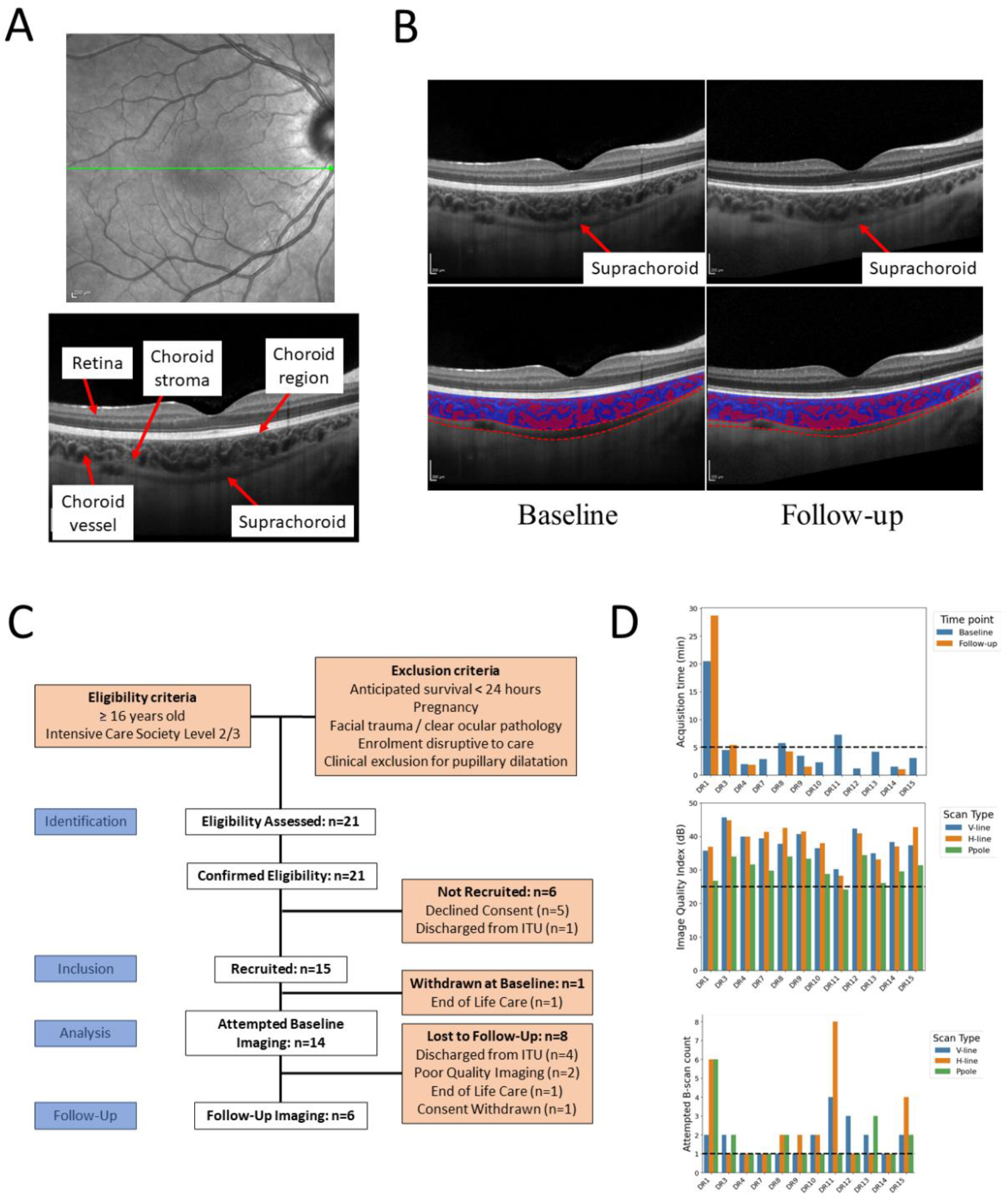
(A) En face retinal scan with location of acquisition in green (top), and optical coherence tomography (OCT) B-scan with landmarks annotated (bottom). (B) Patient with measurable suprachoroid (top), with observed change in suprachoroidal space thickness between time points (bottom). (C) Flowchart summarising patient recruitment. (D) Per-patient metadata from OCT acquisition describing scan time (top), image quality per scan type (middle) and number of scans attempted per visit (bottom).

### Statistical analysis

We measured feasibility in terms acquisition time, image quality index (Heidelberg Q-score),^15^ and number of attempts taken.

The distributions of choroidal and clinical data were reviewed graphically and possible pairwise associations with choroidal metrics were explored using scatterplots. We fitted curves to plots that suggested linear or non-linear associations. Since these associations are exploratory and hypothesis-generating we do not report p-values.

## Results

### Participants

Twenty-one patients were screened and eligible, 15 were recruited, 14 had baseline imaging attempted and 6 also had follow-up imaging (Figure 1C). Demographics of participants who underwent imaging are reported in Table 1.

**Table 1:** Patient summary characteristics at baseline and follow-up.

| Patient Characteristics | Time point |  |  |
| --- | --- | --- | --- |
|  | Baseline | Follow-Up | Laboratory Reference¶ |
| **N (Patients)** | 14 | 6 |  |
| **Age (Mean, Years):** | 56.2±15.3 | 47.0±19.0 | - |
| **Sex (male)** | 10 (71) | 3 (50) | - |
| **Body Mass Index (kg/m<sup>2</sup>):** | 28.4±5.9 | 27.3±7.3 | - |
| **APACHE2\* Score at Critical Care Admission** | 20.4±5.9 | 19±6.6 | - |
| **Days Since Escalation to Level 2 or Level 3 Care\*\*** | 9.1±8.8 | 11.7±11.0 | - |
| **Proportion of Patients Receiving Level 3 Care\*\*** | 14 (100) | 6 (100) | - |
| **Primary Diagnosis on Admission** |  |  |  |
| Cardiac Arrest: | 1 (7) | 0 (0) | - |
| Type 1 Respiratory Failure† | 2 (14) | 2 (34) | - |
| Overdose | 1 (7) | 1 (17) | - |
| Toxic Shock Syndrome | 1 (7) | 1 (17) | - |
| Acute Biliary Disease‡ | 2 (14) | 1 (17) | - |
| Elective Igor-Lewis Oesophagectomy | 2 (14) | 0 (0) | - |
| Elective Whipple's Procedure | 1 (7) | 1 (17) | - |
| Elective Liver Transplantation | 3 (21) | 0 (0) | - |
| Elective Thoraco-aortic Aneurysm Repair | 1 (7) | 0 (0) | - |
| **Indicators of Circulatory Function** |  |  |  |
| Heart Rate (min<sup>-1</sup>) | 87.9±16.9 | 87.8±18.2 | - |
| Mean Arterial Pressure (mmHg) | 84.0±15.3 | 87.6±14.3 | - |
| 24 Hour Fluid Balance (mL) | 460.4±1931.2 | -366.5±560.1 | - |
| Cumulative Fluid Balance Since Treatment Escalation (mL) | 5376.8±8461.5 | 6423±11896.1 | - |
| **Predictors of Circulatory Function** |  |  |  |
| Highest Lactate 24-Hours Preceding Sampling (mmol/L) | 1.54±1.00 | 0.98±0.30 | 0.5-1.6 |
| Creatinine Preceding Sampling (mmol/L) | 123.6±83.3 | 117.2±136.7 | 50-111 |
| Haemoglobin Preceding Sampling (g/L) | 95.2±20.1 | 92.7±21.3 | 115-180 |
| Haematocrit Preceding Sampling | 0.28±0.06 | 0.28±0.06 | 0.36-0.52 |
| Albumin Preceding Sampling (g/L) | 18.3±4.2 | 17.5±3.8 | 36-47 |
| **Receipt of Organ Supportive Therapies** |  |  |  |
| Receipt of Invasive Mechanical Ventilation | 4 (29) | 0 (0) | - |
| FiO<sub>2</sub> Preceding Sampling | 31.0±19.5 | 23.8±5.6 | - |
| SaO<sub>2</sub> Preceding Sampling | 96.7±2.2 | 98.0±1.4 | - |
| Receipt of Vasopressor Therapy | 3 (21) | 1 (17) | - |
| Receipt of Intra-Aortic Balloon Pump | 6 (43) | 2 (33) | - |
| Receipt of Renal Replacement Therapy | 0 (0) | 0 (0) | - |
| Receipt of Intravenous Sedation§ | 3 (21) | 1 (17) | - |
| **Glasgow Coma Scale Preceding Sampling (Median, IQR)** | 15, 3 | 15, 0 | - |
| **Choroid measurements (Median, IQR)** |  |  |  |
| Subfoveal choroid thickness (µm) | 249 (134) | 309 (150) | - |
| Area (mm<sup>2</sup>) | 0.82 (0.62) | 1.15 (0.68) | - |
| Luminal area (mm<sup>2</sup>) | 0.36 (0.35) | 0.60 (0.39) | - |
| Choroidal vascular index | 0.44 (0.05) | 0.52 (0.07) | - |
| **Suprachoroidal measurements (Median, IQR)** |  |  |  |
| Subfoveal suprachoroidal thickness (µm) | 64 (34) | 48 (26) | - |
| Suprachoroidal area (mm<sup>2</sup>) | 0.22 (0.16) | 0.21 (0.13) | - |

**Notes:** All values are N (%) or Mean (standard deviation), unless specified otherwise. \* Acute Physiology and Chronic Health Evaluation Score 2 -(APACHE2); \*\* Intensive Care Society Level 2 (high-dependency) or Level 3 (intensive treatment) care; † Type 1 Respiratory Failure: one case secondary to pneumonia (concomitant bacterial and viral); ‡ Biliary Disease: one case of sepsis secondary to cholangitis, one patient with acute kidney injury following cholecystitis; ¶ Biochemical reference ranges as indicated by local health board (NHS Lothian); § Sedation including either intravenous propofol or clonidine infusion.

### Feasibility

Table 2 and Figure 1D show feasibility data. Baseline imaging was attempted in 14/15 recruited participants (the exception was owing to clinical deterioration and end of life care), and follow-up imaging attempted in six of these. Baseline imaging was successful in 12 of 14 (86%) participants and follow-up imaging was successful in all six attempts (100%). Median (IQR) acquisition time was 2min (0min 48sec to 4min 18sec) (Figure 1D, top). Average (SD) Q-score for horizontal-line scans was 38.9 (4.6) (Figure 1D, middle). Median (IQR) number of scan attempts needed per visit was 1.5 (1 to 2) (Figure 1D, bottom). Details of imaging sessions are in Table 2. The Inability to image two participants was attributed to agitation or abnormal retinal anatomy (high myopia and geographic atrophy).

**Table 2:** Feasibility data for optical coherence tomography in critical care settings.

| Patient ID** | Captured | GCS* | Tropicamide† | Laterality | Position | Time (m:s)‡ | Operator\$ | SCS visible | Case-Specific Notes |
| --- | --- | --- | --- | --- | --- | --- | --- | --- | --- |
| **Patients Imaged at Baseline** |  |  |  |  |  |  |  |  |  |
| 1 | Yes | 4 | Applied | Right | Supine | 20:27 | A | Yes | Sedated, unable to fixate, large suprachoroid, image artefacts. Camera position affected by ventilator in-situ. |
| 2 | No | 11 | Applied | Right | Upright | N/A¶ | A, B | No | Unable to fixate, cooperate, or follow instructions. Camera position affected by ventilator in-situ. |
| 3 | Yes | 15 | Applied | Right | Upright | 04:32 | A | Yes | Required suctioning of airway secretions during protocol. Camera position affected by ventilator in-situ. |
| 4 | Yes | 15 | Not applied§ | Right | Supine | 01:59 | B | Yes |  |
| 5 | No | 3 | Applied | Both | Supine | 13:32 | A | No | Sedated, required saline eye drops for dry eyes. Difficulty locating macula due to previous ophthalmic conditions (AMD). Camera position affected by ventilator in-situ. |
| 7 | Yes | 15 | Applied | Right | Supine | 02:53 | A | Yes | Oxygen face mask in-situ. |
| 8 | Yes | 15 | Applied | Right | Upright | 05:44 | A | Yes | Camera position affected by nebuliser in-situ. Required suctioning mid-protocol. Bedside access blocked ipsilaterally. |
| 9 | Yes | 15 | Applied | Right | Upright | 03:26 | A | Yes | Lack of head support due to upright positioning, lead to difficulty fixating. |
| 10 | Yes | 15 | Applied | Right | Upright | 02:19 | A | Yes | Patient hearing impaired with difficulty fixating. |
| 11 | Yes | 10 | Applied | Right | Supine | 07:15 | A | No | Required saline eye drops for dry eyes. Partially sedated. Highly myopic eyes, large area of peripapillary atrophy. Difficultly fixating. Camera position affected by ventilator in-situ. |
| 12 | Yes | 15 | Applied | Right | Upright | 01:11 | B | No | Poor cooperating and fixation due to patient discomfort. |
| 13 | Yes | 15 | Applied | Right | Upright | 04:09 | A | No | Patient drowsy with difficulty fixating. |
| 14 | Yes | 15 | Applied | Right | Supine | 01:30 | A | Yes |  |
| 15 | Yes | 15 | Applied | Right | Supine | 03:04 | A | Yes | Difficulty imaging deep choroid with enhanced depth OCT. |
| **Patients Imaged at Follow-Up** |  |  |  |  |  |  |  |  |  |
| 1 | Yes | 11 | Applied | Right | Supine | 28:40 | A | Yes | Sedated, unable to fixate, Camera position affected by ventilator in-situ |
| 3 | Yes | 15 | Applied | Right | Supine | 05:27 | A | Yes |  |
| 4 | Yes | 15 | Applied | Right | Supine | 01:52 | A | Yes | Protocol completed despite OCT power supply issues. |
| 8 | Yes | 15 | Applied | Right | Supine | 04:14 | B | Yes | Horizontal imaging repeated to improve capture quality. |
| 9 | Yes | 15 | Not applied§ | Right | Supine | 01:32 | A | Yes | OCT imaging facilitated by supine position. |
| 14 | Yes | 15 | Applied | Right | Upright | 01:04 | A | Yes |  |

**Abbreviations:** ITU, Intensive Therapy Unit (Level 3 care). OCT: optical coherence tomography, SCS: suprachoroidal space, m:s, minutes:seconds. **Notes:** All recruitment and imaging took place in ITU. There were no adverse events from imaging, and no case-specific comments imply imaging was completed without issue. \*\* Patient 6 was excluded before baseline imaging due to clinical deterioration and end of life care; † Oblique Illumination Test (to assess risk of acute closed-angle glaucoma) were negative for those following application of tropicamide mydriatic eye drops; ‡ Time estimated as the difference in acquisition time between first and last scan acquired; \*Glasgow Coma Scale (0-15); § Patient right eye was clinically dilated, with no requirement for mydriasis. ¶ Only one scan was collected so timing could not be estimated; \$ Operators A were authors C.H., J.B. and B were authors E.G., J.B.

### Choroidal variation

Baseline choroidal thickness, CVI and suprachoroidal space thickness are shown in Figure 2A. The suprachoroidal space was visible in nine (75%) of 12 participants, in some cases was markedly enlarged (Figure 1B), with the interquartile range notably larger than previously reported in a healthy cohort,^16^ (Figure 2A, bottom shaded blue).

**Figure 2:**
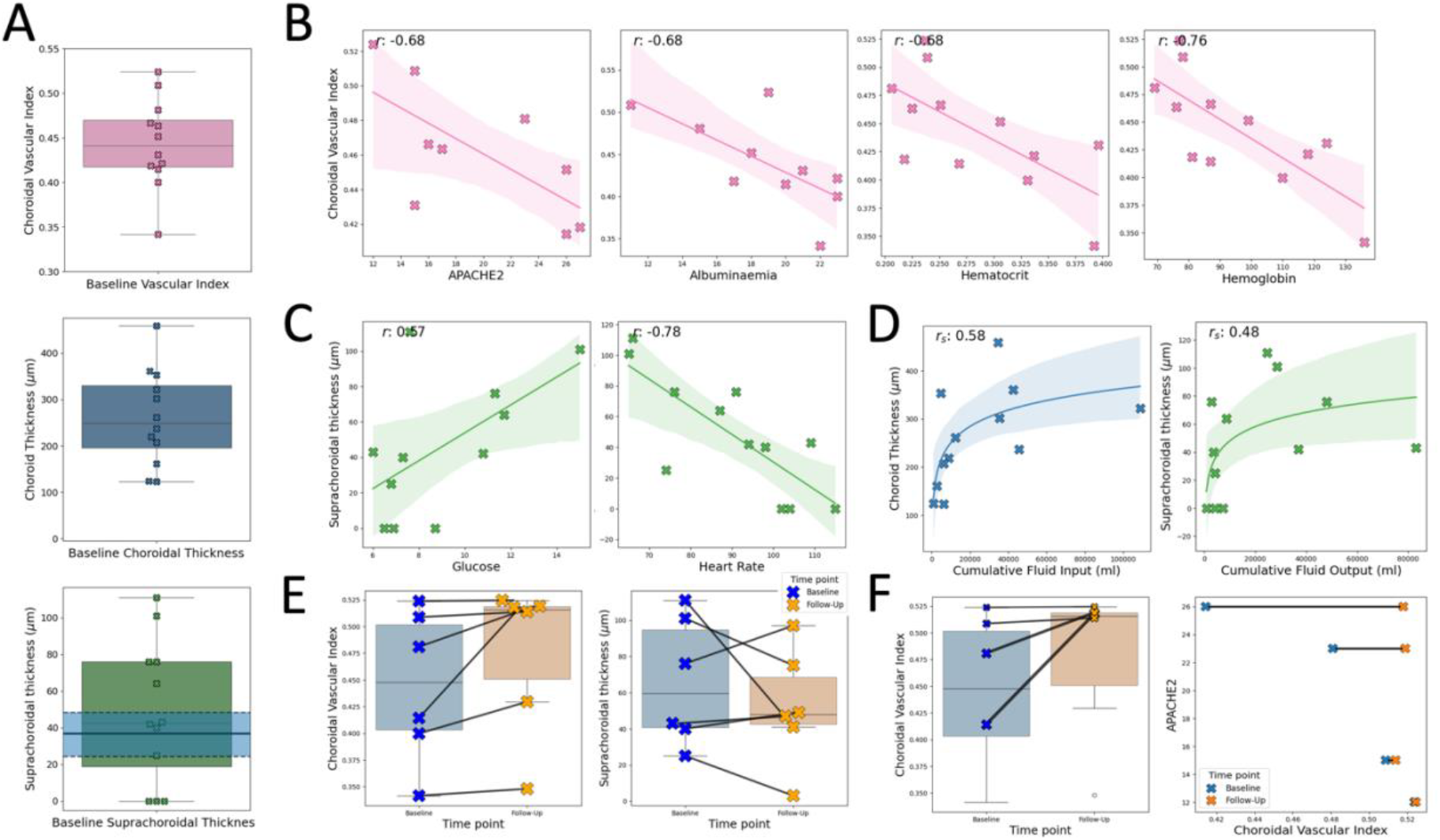
(A) Choroidal variation at baseline for CVI (top), subfoveal thickness (middle) and suprachoroidal thickness (bottom), with shaded blue region representing median and IQR from Yiu, et al.,^16^ (B) Observed trends between baseline CVI and APACHE2, Albuminaemia (g/L), Haematocrit (%) and Haemoglobin (g/L). (C) Observed trends between suprachoroidal thickness with Glucose (mmol/L) and Heart rate (bpm). (D) Observed, non-linear and monotonic trends between choroid thickness and cumulative fluid input (ml), and suprachoroidal thickness and cumulative fluid output (ml). (E) Observed change in CVI and suprachoroidal thickness between baseline and follow-up time points. (F) Magnitude of CVI change appeared to increase as a function of APACHE2 score (2 patients of the 6 who were followed-up did not have an APACHE2 score).

We found that increased haematological markers and disease severity significantly associated with a thinner choroid, reduced choroid vessel density and increased suprachoroidal space. Specifically, baseline choroid vessel density (CVI) was lower in patients with a greater Acute Physiology and Chronic Health Evaluation (APACHE) II score (r= –0.68), albuminaemia (r=−0.68), haematocrit (r= –0.68) and haemoglobin (r= –0.76), patients with thinner choroids had increased haemoglobin (r –0.57), and the suprachoroidal space tended to be smaller in patients with lower glucose (r= +0.57) and higher heart rates (Figure 2C). A full list of pairwise comparisons with pearson/spearman correlations between choroidal measurements and clinical measures are shown in Supplement 4.

We also found that patients with increased cumulative fluid levels had thicker choroids with increased vascularity (CVI) and with larger suprachoroidal spaces. The increase in these measurements was monotonic but non-linear (spearman s for cumulative input fluid: choroidal thickness, s=+0.58; suprachoroidal space thickness s=+0.47; CVI, s=+0.57; Spearman s for cumulative output fluid: choroidal thickness, s=+0.58; suprachoroidal space thickness, s=+0.48; CVI s=+0.55). Some notable examples are shown in Figure 2D.

There was no evidence of association between choroidal measurements with systemic blood pressure, nor between haematological markers and systemic blood pressure (mean arterial blood pressure and haemoglobin r=+0.02, haematocrit r=−0.08 and albuminaemia r=−0.03).

In the six participants with follow-up imaging, CVI and suprachoroidal thickness changed substantially in some individuals (Figure 2E). The largest increases in CVI appeared to be in patients with the highest APACHE II scores (Figure 2F).

## Discussion

We found that EDI-OCT imaging of the choroid is feasible in patients with shock in ITU/HDU and reveals substantial variation in the size and vascularity (CVI) of the choroid in this context. Our exploratory results suggest that choroidal measurements may reflect systemic fluid status and rheology, and that EDI-OCT is sensitive to changes within individuals over time.

These observations are of particular interest given the absence of any notable relationship between choroidal measurements and macrovascular markers of systemic blood pressure, and similarly between rheological markers and systemic blood pressure. This may suggest the choroidal vascular bed is more informative at reflecting the microvascular environment than macro-scale physiological measurements and could be a useful sampling site for assessing organ perfusion in the context of critical illness.

We also found that the suprachoroidal space can be markedly enlarged in this patient group. This frequency (75%) and magnitude of suprachoroidal space we found is unusual. A thin suprachoroidal space may be visible in up to 44% of healthy people aged 55-85 years,^16^ but an obviously visible space is usually associated with ocular disease such as severely low eye pressure or retinochoroidal inflammation.^17^

To the best of our knowledge, this is the first study to report choroidal imaging in this setting, and across multiple diagnoses. A few studies have reported retinal imaging in ITU,^6,9^ or after discharge from ITU.^18,19^ Liu et al.,^9^ demonstrated OCT and OCT-Angiography (OCT-A) feasibility in a similar setting, and our study extends this by investigating a larger cohort and suggesting the feasibility of enhanced depth imaging OCT of the choroid. Additionally, Courtie et al.,^6^ found changes in retinal blood flow using OCT-A in patients undergoing elective oesophagostomy, supporting our findings of abnormal intra-ocular anatomy in critical illness. Measurements of retina and choroid, and particularly change over time within individuals, may help evaluate the degree of microcirculatory dysfunction. Compared to sublingual sidestream-dark field microscopy,^20^ OCT has the advantage of precise spatial registration of images across time and is therefore able to detect micron-scale changes.

Our study had a small sample and did not follow-up participants after discharge from ITU. These limitations did not impact our main aim (assessing feasibility), but future studies designed to test hypotheses about associations between choroidal metrics and systemic physiology should follow patients up after discharge. Importantly, while we demonstrate its feasibility, effective imaging with the OCT Spectralis Flex required two or three operators and co-operation of departmental research nurses and the wider, direct clinical care team. However, the size, cost, and ease of use of portable OCT equipment is likely to improve in the future with advancements in the technology– creating the potential for retinal microcirculatory biomarkers of systemic disease,^21,22^ especially since retinochoroidal OCT images can now be analysed with open-source computational methods.^12–14,23^

## Data Availability

The data that support the findings of this study are not openly available due to reasons of sensitivity and are available from the corresponding author upon reasonable request.
Software used for image analysis are three image segmentation algorithms, DeepGPET, GPET and MMCQ, used for choroid segmentation in optical coherence tomography images. See links below to their open-access codebases.

https://github.com/jaburke166/deepgpet

https://github.com/jaburke166/gaussian_process_edge_trace

https://github.com/jaburke166/mmcq

## Acknowledgements

The authors would like to thank all participants in the study as well as all staff in the ITU ward at the Royal Infirmary of Edinburgh who contributed to data collection and image acquisition for this study.

## Ethics approval and consent to participate

The study involves human participants and was approved by Lothian NHS Board (REC 22//SS//0055), Lothian R&D Project No 2022/0190. Participants (or next of kin otherwise) gave informed consent to participate in the study before taking part and the declaration of Helsinki was followed throughout.

## Author’s contributions

GC: Project administration, data collection, formal analysis, writing – original draft, review & editing.

JB: Data collection, software, formal analysis, writing – original draft, review & editing.

CH: Data collection.

EG: Data collection.

ND: Supervision.

SK: Supervision.

TM: Supervision.

KB: Supervision, writing – review & editing.

DG: Supervision, writing – review & editing.

IM: Supervision, writing – original draft, review & editing.

## Availability of data and materials

The data that support the findings of this study are not openly available due to reasons of sensitivity and are available from the corresponding author upon reasonable request. Data are located in controlled access data storage at the University of Edinburgh.

Software used for image analysis are three image segmentation algorithms, DeepGPET, GPET and MMCQ, used for choroid segmentation in optical coherence tomography images. These are listed below:

Project name: DeepGPET

Project home page: https://github.com/jaburke166/deepgpet

Archived version: https://doi.org/10.1167/tvst.12.11.27

Operating system(s): Platform independent

Programming language: Python

Other requirements: None

License: GNU GPL (CC-BY)

Project name: Gaussian Process Edge Tracing

Project home page: https://github.com/jaburke166/gaussian_process_edge_trace

Archived version: https://doi.org/10.1109/TIP.2021.3128329

Operating system(s): Platform independent

Programming language: Python

Other requirements: None

License: GNU GPL (CC-BY)

Project name: MMCQ

Project home page: https://github.com/jaburke166/mmcq

Archived version: https://doi.org/10.1167/iovs.65.6.6

Operating system(s): Platform independent

Programming language: Python

Other requirements: None

License: GNU GPL (CC-BY)

## References

1. Sakr Y, Gath V, Oishi J, et al. Characterization of buccal microvascular response in patients with septic shock. Eur J Anaesthesiol. 2010;27(4):388–394. doi:10.1097/EJA.0b013e3283349db3

2. Gazmuri RJ, de Gomez CA. From a pressure-guided to a perfusion-centered resuscitation strategy in septic shock: Critical literature review and illustrative case. J Crit Care. 2020;56:294–304. doi:10.1016/j.jcrc.2019.11.008

3. null null. Early, Goal-Directed Therapy for Septic Shock — A Patient-Level Meta-Analysis. New England Journal of Medicine. 2017;376(23):2223–2234. doi:10.1056/NEJMoa1701380

4. Maitland Kathryn, Kiguli Sarah, Opoka Robert O., et al. Mortality after Fluid Bolus in African Children with Severe Infection. New England Journal of Medicine. 2011;364(26):2483–2495. doi:10.1056/NEJMoa1101549

5. Hutchings SD, Naumann DN, Hopkins P, et al. Microcirculatory Impairment Is Associated With Multiple Organ Dysfunction Following Traumatic Hemorrhagic Shock: The MICROSHOCK Study. Critical Care Medicine. 2018;46(9):e889. doi:10.1097/CCM.0000000000003275

6. Courtie E, Veenith T, Logan A, Denniston AK, Blanch RJ. Retinal blood flow in critical illness and systemic disease: a review. Annals of Intensive Care. 2020;10(1):152. doi:10.1186/s13613-020-00768-3

7. MacCormick IJC, Beare NAV, Taylor TE, et al. Cerebral malaria in children: using the retina to study the brain. Brain. 2014;137(Pt 8):2119–2142. doi:10.1093/brain/awu001

8. Farrah TE, Dhillon B, Keane PA, Webb DJ, Dhaun N. The eye, the kidney, and cardiovascular disease: old concepts, better tools, and new horizons. Kidney Int. 2020;98(2):323–342. doi:10.1016/j.kint.2020.01.039

9. Liu X, Kale AU, Capewell N, et al. Optical coherence tomography (OCT) in unconscious and systemically unwell patients using a mobile OCT device: a pilot study. BMJ Open. 2019;9(11):e030882. doi:10.1136/bmjopen-2019-030882

10. Reiner A, Fitzgerald MEC, Del Mar N, Li C. Neural control of choroidal blood flow. Prog Retin Eye Res. 2018;64:96–130. doi:10.1016/j.preteyeres.2017.12.001

11. Kuriakose T. The Slit Lamp Examination. In: Kuriakose T, ed. Clinical Insights and Examination Techniques in Ophthalmology. Springer; 2020:55–62. doi:10.1007/978-981-15-2890-3_6

12. Burke J, Engelmann J, Hamid C, et al. An Open-Source Deep Learning Algorithm for Efficient and Fully Automatic Analysis of the Choroid in Optical Coherence Tomography. Translational Vision Science & Technology. 2023;12(11):27. doi:10.1167/tvst.12.11.27

13. Burke J, King S. Edge Tracing Using Gaussian Process Regression. IEEE Transactions on Image Processing. 2022;31:138–148. doi:10.1109/TIP.2021.3128329

14. Engelmann J, Burke J, Hamid C, et al. Choroidalyzer: An Open-Source, End-to-End Pipeline for Choroidal Analysis in Optical Coherence Tomography. Investigative Ophthalmology & Visual Science. 2024;65(6):6. doi:10.1167/iovs.65.6.6

15. Engineering H. SPECTRALIS Product Family User Manual Sw Ver 7.0. Version 7.; 2022.

16. Yiu G, Pecen P, Sarin N, et al. Characterization of the Choroid-Scleral Junction and Suprachoroidal Layer in Healthy Individuals on Enhanced-Depth Imaging Optical Coherence Tomography. JAMA Ophthalmology. 2014;132(2):174–181. doi:10.1001/jamaophthalmol.2013.7288

17. Sadda SR, Schachat AP, Wilkinson CP, et al. Ryan’s Retina E-Book. Elsevier Health Sciences; 2022.

18. Icoz M, Ar ıkan YM, Bayhan GI, Gurturk ISG, Yahsi A. Assessment of the Choroidal Vascular Structure in Multisystem Inflammatory Syndrome in Children. Journal of Pediatric Ophthalmology & Strabismus. 2024;61(2):120–126. doi:10.3928/01913913-20230809-01

19. Gül FC, Timurkaan ES. Evaluation of choroidal thickness with OCT in COVID-19 patients with high D-dimer levels. Sci Rep. 2022;12(1):16826. doi:10.1038/s41598-022-21579-5

20. Bruno RR, Wollborn J, Fengler K, et al. Direct assessment of microcirculation in shock: a randomized-controlled multicenter study. Intensive Care Med. 2023;49(6):645–655. doi:10.1007/s00134-023-07098-5

21. Chopra R, Wagner SK, Keane PA. Optical coherence tomography in the 2020s—outside the eye clinic. Eye. 2021;35(1):236–243. doi:10.1038/s41433-020-01263-6

22. Song G, Jelly ET, Chu KK, Kendall WY, Wax A. A review of low-cost and portable optical coherence tomography. Prog Biomed Eng. 2021;3(3):032002. doi:10.1088/2516-1091/abfeb7

23. Burke J, Engelmann J, Gibbon S, et al. OCTolyzer: Fully automatic toolkit for segmentation and feature extracting in optical coherence tomography and scanning laser ophthalmoscopy data. Published online January 13, 2025. doi:10.48550/arXiv.2407.14128

