## Supplement 1 for "Direct Retinal Imaging for Shock Resuscitation in Critical Ill Adults II (D-RISC II)"

### Supplementary Material 1: OCT Imaging Protocol

Optical coherence tomography scans were collected using the spectral domain Heidelberg SPECTRALIS Flex module (Heidelberg Engineering, Heidelberg, Germany). All scans collected were macula-centred with a 30 degrees (approximately 9 $mm^2$) field of view. Enhanced depth imaging was activated in order to improve visualisation of the choroid-scleral junction and vasculature. Active eye tracking and automatic real time (ART, the number of B-scans averaged at the same location) was used to help improve image quality, longitudinal registration and reduce speckle noise. Our imaging protocol is enumerated below in order of image capture:

1. A fovea-centred, posterior pole horizontal-line B-scan with an ART of 100
2. A fovea-centred, posterior pole vertical-line B-scan with an ART of 100.
3. A fovea-centred posterior pole volume scan, consisting of equally spaced B-scans, approximately 240 microns apart using an ART of 9. Dependent upon patient cooperation, the field of view for this volume scan was reduced:
   a. Full cooperation: 31 B-scans covering an 8.0 x 6.6 mm region of interest;
   b. Adequate cooperation: 25 B-scans covering an 8.0 x 5.3 mm region of interest;
   c. Challenging cooperation: 25 B-scans a 5.3 x 5.3 mm region of interest.
4. If the patient was still fully cooperative, a circular peripapillary B-scan centred on the optic nerve head using an ART of 100 was taken.
