## Supplement 2 for "Direct Retinal Imaging for Shock Resuscitation in Critical Ill Adults II (D-RISC II)"

### Supplementary Material 2: Clinical Variables Collected

**Table 1:** Clinical and ocular variables collected through paper-based report form.

| Variable Name | Units | Measure | Source | Baseline | Follow-Up |
|---|---|---|---|---|---|
| Age | Years | Independent | CRF | Yes | No |
| Sex (Male/Female) | Binary | Independent | CRF | Yes | No |
| Height | cm | Independent | CRF | Yes | No |
| Weight | kg | Independent | CRF | Yes | No |
| BMI | N/A | Independent | CRF | Yes | No |
| APACHE2 | N/A | Independent | CRF | Yes | No |
| Setting (ITU/HDU) | Binary | Independent | CRF | Yes | No |
| Diagnosis | Free | Independent | CRF | Yes | No |
| CCI | N/A | Independent | CRF | Yes | No |
| SARS-CoV-2 Status | Binary | Independent | CRF | Yes | No |
| Previous Ophthalmology | Free | Independent | CRF | Yes | No |
| Additional Comorbidities | Free | Independent | CRF | Yes | No |
| Hospital Admission | Days | Independent | CRF | Yes | Yes |
| ITU Admission | Days | Independent | CRF | Yes | Yes |
| Eye Imaged | Chiral | Process | CRF | Yes | Yes |
| Flashlight Test Performed | Binary | Process | CRF | Yes | Yes |
| Flashlight Test Result | Binary | Process | CRF | Yes | Yes |
| Tropicamide Applied | Binary | Process | CRF | Yes | Yes |
| Patient Position | Free | Process | CRF | Yes | Yes |
| Image Time | D-T | Process | CRF | Yes | Yes |
| Acquisition time | T | Process | CRF | Yes | Yes |
| Image quality | Q | Process | CRF | Yes | Yes |
| Scans attempted | N | Process | CRF | Yes | Yes |
| Saline (Tears) Applied | Binary | Process | CRF | Yes | Yes |
| Image Adverse Events | Free | Process | CRF | Yes | Yes |
| Imaging Challenges | Free | Process | CRF | Yes | Yes |
| Patient Airway | Free | Independent | CRF | Yes | Yes |
| Respiratory Therapy | Free | Independent | CRF | Yes | Yes |
| Fraction Inspired Oxygen | % | Independent | CRF | Yes | Yes |
| PEEP | $cmH_2O$ | Independent | CRF | Yes | Yes |
| Vasopressor Required | Binary | Independent | CRF | Yes | Yes |
| Invasive Blood Pressure | mmHg | Independent | CRF | Yes | Yes |
| VAECMO Required | Binary | Independent | CRF | Yes | Yes |
| RRT Required | Binary | Independent | CRF | Yes | Yes |
| Sedation Infusion | mcg/hr | Independent | CRF | Yes | Yes |
| Vasopressor Infusion | mcg/hr | Independent | CRF | Yes | Yes |
| Analgesia Infusion | mcg/hr | Independent | CRF | Yes | Yes |
| 24 Hour Fluid Input | ml | Independent | CRF | Yes | Yes |
| 24 Hour Fluid Output | ml | Independent | CRF | Yes | Yes |
| 24 Hour Fluid Balance | ml | Independent | CRF | Yes | Yes |
| Cumulative Fluid Input | ml | Independent | CRF | Yes | Yes |
| Cumulative Fluid Output | ml | Independent | CRF | Yes | Yes |
| Cumulative Fluid Balance | ml | Independent | CRF | Yes | Yes |
| Peripheral Pulse Oximetry | % | Independent | CRF | Yes | Yes |
| Respiratory Rate | bpm | Independent | CRF | Yes | Yes |
| Systolic Blood Pressure | mmHg | Independent | CRF | Yes | Yes |
| Diastolic Blood Pressure | mmHg | Independent | CRF | Yes | Yes |
| Peripheral Capillary Refill Time | seconds | Independent | CRF | Yes | Yes |
| Glasgow Coma Score | N/A | Independent | CRF | Yes | Yes |
| Peripheral Temperature | Celsius | Independent | CRF | Yes | Yes |
| Right Heart Catheterisation | $cmH_2O$ | Independent | CRF | Yes | Yes |
| Haemocrit | % | Independent | CRF | Yes | Yes |
| Haemoglobin | g/L | Independent | CRF | Yes | Yes |
| Albumin | g/L | Independent | CRF | Yes | Yes |

| | | | | | |
|---|---|---|---|---|---|
| Creatinine | mmol/L | Independent | CRF | Yes | Yes |
| Heart Rate | bpm | Independent | CRF | Yes | Yes |
| Glucose | mmol/L | Independent | CRF | Yes | Yes |
| Bicarbonate | mmol/L | Independent | CRF | Yes | Yes |
| Highest Lactate (24 Hour) | mmol/L | Independent | CRF | Yes | Yes |
| Highest C-Reactive Protein (24 Hour) | mmol/L | Independent | CRF | Yes | Yes |

**Abbreviations:** CRF, Case Report Form; BMI, Body Mass Index; APACHE2, Acute Physiological and Chronic Health Evaluation;  ITU, Intensive Therapy Unit, HDU, High Dependency Unity; CCI, Charlson Comorbidity Index; N/A, Not Applicable; D-T, Date-Time; PEEP, Positive End Expiratory Ventilation VAECMO, Veno-Arterial Extracorporeal Membrane Oxygenation; RRT, Renal Replacement Therapy; Q, Q-score signal-to-noise ratio from imaging device; T, Time; N, whole number.
