## Supplement 3 for "Direct Retinal Imaging for Shock Resuscitation in Critical Ill Adults II (D-RISC II)"

### Supplementary Material 3: OCT B-scan imaging and choroid measurements

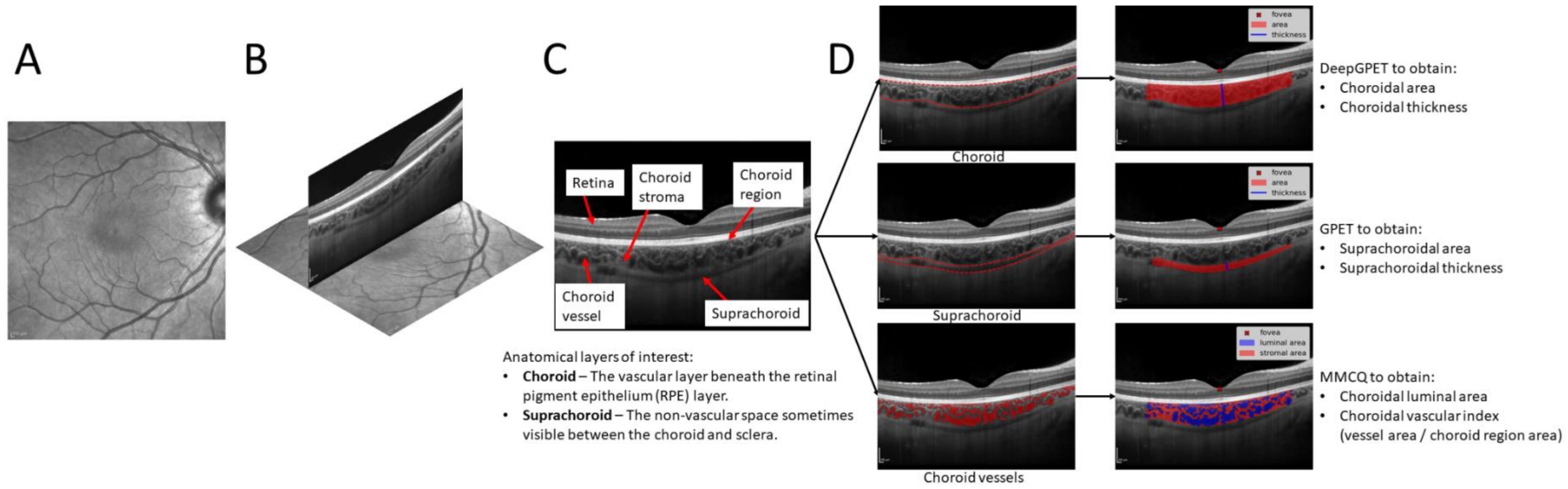


**Figure 1:** Diagram of choroidal measurements of interest for this study. (A) En face fundus image of the retinal vasculature. (B) OCT B-scan overlaid onto the fundus image at the location of acquisition. (C) OCT B-scan with annotations of relevant intra-ocular structures. (D) Automatic segmentations of the choroid region (using DeepGPET), vessels (using MMCQ) and suprachoroid (using GPET) with corresponding measurements of interests overlaid.

For each OCT B-scan, choroidal measurements of subfoveal choroid thickness, area, vessel area and vascular index were taken using a fovea-centred region of interest (ROI). Thickness was measured as a straight-line micron distance from upper-to-lower choroid boundary, drawn perpendicular to the upper boundary to account for any choroidal curvature. Choroid area and vessel area was measured as the number of pixels contained within the ROI, converted into $mm^2$. Choroid vascular index is a dimensionless metric which measures the proportion of vessel pixels to choroid pixels. Figure 1 shows an en face retinal scan (A), an exemplar fovea-centred OCT B-scan and its colocation on the retinal scan (B), the B-scan with landmarks of interest annotated (C), and choroidal segmentations and measurements overlaid (D).
