## Supplement 4 for "Direct Retinal Imaging for Shock Resuscitation in Critical Ill Adults II (D-RISC II)"

**Supplementary Material 4: Correlation heatmap between choroid and clinical measures**

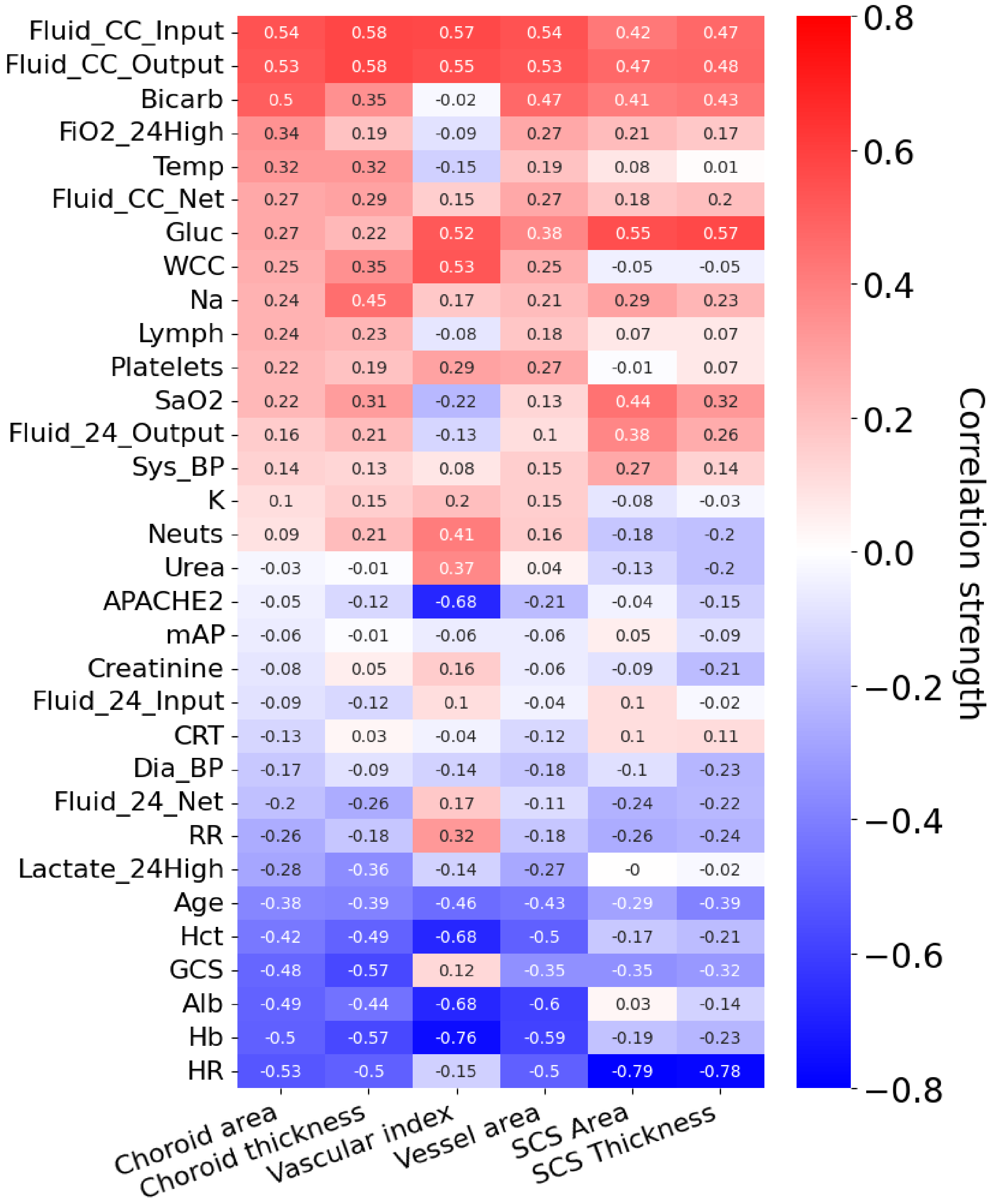


**Figure 1**: Correlation heatmap between choroidal measurements (horizontal axis) and clinical measurements (vertical axis), with correlation coefficients superimposed and ordered by choroidal area (far left). **Notes:** All correlations are Pearson correlation coefficients, with exception of cumulative fluid input/output/net which use the Spearman correlation coefficient due to the observed non-linear trend. **Abbreviations:** SCS, suprachoroidal space; CC, cumulative; Gluc, glucose; CRT, capillary refill time; CRP, C-reactive protein; FiO2, fraction inspired oxygen; WCC, white cell count; RR, respiratory rate; GCS, Glasgow coma scale; Hct, haematocrit; Hb, haemoglobin; HR, heart rate; Alb, albuminuria; Temp, peripheral temperature; Bicard, bicarbonate; APACHE2, Acute Physiology and Chronic Health Evaluation Score 2; K, potassium; mAP, mean arterial pressure; Na, sodium; Dia_BP, diastolic blood pressure; Sys_BP, systolic blood pressure; Neuts, Neutrophils; Sa02, oxygen saturation.
